# Discordance between HbA_1C_ and Glucose Tests for the Diagnosis of Prediabetes in a Filipino-American Cohort

**DOI:** 10.64898/2026.01.09.26343777

**Authors:** Maria Elizabeth P. Mercado, Sarah Kim, Eva Maria Cutiongco-de la Paz, Mark Seielstad, Elizabeth Paz-Pacheco, Elizabeth J. Murphy

## Abstract

**Introduction:** The diagnostic utility of HbA1c for identifying prediabetes remains uncertain, particularly across diverse racial and ethnic populations. We evaluated concordance between HbA1c, fasting plasma glucose (FPG), and oral glucose tolerance testing (OGTT) in a Filipino-American cohort.

**Research Design and Methods:** We analyzed cross-sectional data from 149 Filipino-American adults without known diabetes in the San Francisco Bay Area. Prediabetes prevalence was assessed using American Diabetes Association criteria for HbA1c, FPG, and OGTT. Agreement between diagnostic methods was evaluated, and metabolic characteristics were compared across glycemic categories.

**Results:** Prediabetes prevalence differed substantially by diagnostic method: 35% by HbA1c, 8.1% by FPG, and 8.7% by OGTT. Overall agreement between tests was low. Among individuals classified as prediabetic by HbA1c, the majority did not meet criteria by FPG or OGTT. However, higher HbA1c levels were associated with increased body mass index, waist-hip ratio, blood pressure, triglycerides, fasting insulin, and HOMA-IR.

**Conclusions:** In this Filipino-American cohort, HbA1c identifies a substantially larger group of individuals with prediabetes compared to glucose-based tests, with limited concordance between methods. Individuals classified as prediabetic by HbA1c exhibit features consistent with insulin resistance. These findings highlight potential differences in metabolic phenotypes captured by alternative diagnostic criteria and underscore the need for longitudinal studies to determine their relative prognostic value.

**What is already known on this topic:** HbA1c, FPG, and OGTT are all used to diagnose prediabetes, but prior studies show poor agreement between these tests and substantial variation in prevalence estimates across populations. Evidence suggests these differences may be influenced by underlying physiology and race/ethnicity, yet data in Filipino-Americans—especially for prediabetes—have been limited.

**What this study adds:** In Filipino-Americans, HbA1c identifies a much higher prevalence of prediabetes than FPG or OGTT, with low concordance between methods. Individuals classified as prediabetic by HbA1c alone show metabolic features consistent with insulin resistance, suggesting this group represents a meaningful at-risk phenotype.

**How this study might affect research, practice or policy:** These findings suggest that reliance on a single diagnostic test may misclassify risk and that HbA1c may capture additional at-risk individuals in this population. They support the need for longitudinal studies and potential reconsideration of diagnostic thresholds or combined testing approaches tailored to specific populations.

## Introduction

The key to successful diabetes prevention is identifying those at highest risk of developing diabetes to allow for therapeutic intervention. Since the introduction of glycated hemoglobin (HbA_1C_) for the diagnosis of diabetes mellitus (DM) and prediabetes in 2010 by the American Diabetes Association (ADA)^1^, there has been ongoing debate about the diagnostic accuracy of this measure^2–10^ Although both the World Health Organization (WHO)^11^ and (ADA)^12^ agree on the use of HbA_1C_ for the diagnosis of DM, they are not in agreement for its use in prediabetes.

Overall, agreement between glycated hemoglobin (HbA_1C_) and fasting plasma glucose (FPG) and/or 2-hour oral glucose tolerance test (OGTT) for diagnosing prediabetes has been weak ^13–16^ with highly variable prevalence for prediabetes. Fasting plasma glucose, OGTT and HbA_1C_ measure distinctly different processes with presumably heterogenous underlying pathophysiology. Broadly speaking, FPG is more reflective of hepatic insulin resistance while OGTT is more reflective of peripheral insulin resistance. HbA_1C_ will reflect a combination of the two. Thus, it should not be surprising that large population studies show discordance between these measures. Studies have also shown significant discordance across racial and ethnic groups. NHANES data show the relationship between HbA_1C_ and FPG, and between HbA_1C_ and OGTT are curvilinear and vary based on race and ethnicity^10^ suggesting the selection of a single cut-off point for the diagnosis of diabetes and prediabetes is difficult. Studies of Mexican American,^7^ Japanese,^18,19^ and South Asian^20^ populations have shown a lower prevalence of prediabetes using HbA_1C_ compared to FPG and/or OGTT. In contrast, data from Chinese^13^ (especially those with obesity), high metabolic risk Hispanic^14^, and Palestinian Arab^15^ cohorts show a higher prevalence of prediabetes using HbA_1C_ compared to FPG or OGTT. In those populations, a significant percentage of individuals identified as having prediabetes by HbA_1C_ were found to have normal FPG ^13–15^ or normal OGTT ^13–16^. Recent literature suggests varying cut-offs for diagnosis using HbA1C based on race and ethnicity, which implies the need to explore HbA1C’s performance in varied populations.

Among a Filipino-American cohort, the prevalence of diabetes was higher with OGTT than by either HbA_1C_ or FPG^22^. However, no study has yet explored the performance of HbA_1C_ in the prediabetes range. Using data from the Filipino Health Study, we examined the relationship between FPG, OGTT and HbA_1C_ in diagnosing prediabetes in Filipino-Americans. We also explored markers of insulin resistance and metabolic syndrome in subgroups with the ultimate aim of assessing the utility of HbA_1C_ in the diagnosis of prediabetes in this population.

## Materials and Methods

### Study participant recruitment

Inclusion criteria for the Filipino Health study were subjects 18 years and older with four grandparents of Filipino descent living in the United States for at least one year at the time of recruitment. Subjects were recruited from the San Francisco Bay area. Exclusion criteria were antibiotic use in the past month (the parent study explores gut microbiome), use of any diabetes medication, current pregnancy, current cancer, and US residence of < 1 year, and MCV < 70 fL as a low MCV is concerning for a possible hemoglobinopathy which could interfere with the HbA_1C_ results. At enrollment a self-reported history was obtained collecting data on medication use, self -reported diabetes status and past medical history. Laboratory studies were obtained the same day. The study protocol was approved by the UCSF Institutional Review Board reference number 16-18705. All participants provided written informed consent prior to enrolment.

### Anthropometric measurements

Participants were weighed on a digital scale (Seca Robusta 813). Height was determined using a stadiometer (Seca 213). Waist and hip circumference were taken following WHO protocol^23^ and were recorded to the nearest 0.5 cm.

Blood pressure was the average of three determinations while sitting using a digital blood pressure monitor (Omron HEM 705 CP).

### Laboratory Evaluation

Blood was collected after a 10 hour fast for CBC, FPG, insulin, lipid panel and HbA_1C_ followed by a 75-g OGTT (EasyDex 10 oz. 75g) with a 2-hour blood glucose. Fasting lipid panel, glucose, insulin and HbA_1C_ measurements were performed by LabCorp (Burlington, North Carolina). Analysis of serum insulin was performed using a 2-site electrochemiluminescent immunoassay on the Roche automated platform (reference interval 2.6 – 24.9 µIU/mL). Analysis of HbA_1C_ was done using the Tina-quant Hemoglobin A1c Gen.3 machine (Roche c513 CTS) with references standardized to DCCT/NGSP.

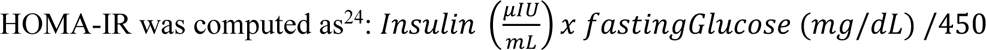

Glycemic categories were based on ADA definitions^12^: normal (FPG < 5.6 mmol/L, OGTT < 7.8 mmol/L, HbA_1C_ < 39 mmol/mol (5.7%)), prediabetes (FPG 5.6 – 6.9 mmol/L, OGTT 7.8 – 11 mmol/L, HbA_1C_ 39 – 46 mmol/mol (5.7% - 6.4%)) and DM (FPG ≥ 7 mmol/L, OGTT ≥ 11.10 mmol/L, HbA_1C_ ≥ 48 mmol/mol (6.5%)). Participants are also grouped into normoglycemia (FPG < 5.6 mmol/L or OGTT < 7.8 mmol/L) and “dysglycemia” (FPG ≥ 5.6 mmol/L or OGTT ≥ 7.8 mmol/L), when applicable. The latter group included subjects who met the criteria for diabetes.

### Statistical Analysis

Differences between tests for dysglycemia were analyzed using independent t-test, one-way analysis of variance, Fisher’s exact, and Kruskal-Wallis as appropriate. Data are expressed as means ± standard deviation for continuous variables and frequencies (percentage) for categorical variables.

Assessment of the relationship between HbA_1C_ and FPG, and between HbA_1C_ and OGTT were modelled following the model by Guo et al.^7^ The models were each assessed for linearity. Outliers were defined as DFBETA values > 0.3 based on plotted DFBETA values. Linearity for the model assessing relationship between HbA_1C_ and FPG did not change with exclusion of outlier points and the full data set was used for unadjusted and adjusted analysis. However, the model assessing the relationship between HbA_1C_ and OGTT suggested nonlinearity which on reassessment was no longer present after excluding outlier points, hence a simple linear model excluding outlier points (n = 143) was used for the final model. Age, gender and BMI were included in the multilinear regression model for the adjusted analyses. Analyses were done using STATA/IC 15.1 (Statacorp, Texas USA).

## Results

### Demographics

A total of 167 Filipino-Americans without previously diagnosed diabetes were enrolled from November 2016 to December 2017. Twelve of those individuals were missing oral glucose tolerance tests and were excluded from this analysis. Another 6 subjects were excluded due to an MCV < 70 fL.

The final cohort (n = 149) was majority female (56%) with an average BMI of 25.7 ± 3.8 kg/m^2^ and age of 42 ± 19 years (Table 1). The majority of subjects had family history of DM (67%) and 35% had hypertension. Another 7% had a history of cardiovascular disease. On average, the cohort was insulin resistant based on an average HOMA-IR of 2.1. Nineteen subjects were taking a statin.

**Table 1:** Characteristics of Filipino-American participants. Data are reported as mean ± standard deviation or n (%), where applicable. BMI - body mass index; FPG – fasting plasma glucose; OGTT – oral glucose tolerance test; MCV – mean corpuscular volume.

|  | **Estimate** | **Range** |
| --- | --- | --- |
| **Age (years)** | 42 ± 19 | 18 – 91 |
| **Female** | 84 (56) |  |
| **Family History of DM** | 100 (67) |  |
| **History of Hypertension** | 71 (35) |  |
| **History of Cardiovascular Disease** | 10 (7) |  |
| ***Anthropometrics*** |  |  |
| **BMI, (kg/m<sup>2</sup>)** | 25.7 ± 3.8 | 17.4 – 38.7 |
| **Waist-Hip-Ratio** | 0.87 ± 0.07 | 0.69 – 1.16 |
| **Waist Circumference (cm)** |  |  |
| *Female* | 81.3 ± 11.0 | 58 – 119 |
| *Male* | 89.8 ± 10.1 | 70 – 114 |
| **Blood Pressure (mmHg)** |  |  |
| *Systolic* | 126 ± 19 | 94 – 184 |
| *Diastolic* | 75 ± 10 | 53 – 100 |
| ***Laboratory Results*** |  |  |
| **Oral Glucose Tolerance Test (mmol/L)** |  |  |
| *0 Hour* | 4.8 ± 0.6 | 3.6 – 6.8 |
| *2<sup>nd</sup> Hour* | 5.5 ± 1.8 | 1.7 – 12.8 |
| **HbA<sub>1C</sub> (%)** | 5.6 ± 0.4 | 4.5 – 7.5 |
| *HbA<sub>1C</sub> (mmol/mol)* | 0.3 ± 0.02 | 0.3 – 0.4 |
| **Insulin (pmol/L)** | 73.6 ± 64.6 | 3.5 – 499.4 |
| **HOMA-IR** | 2.1 ± 2.1 | 0.1 – 18.2 |
| **Lipid Profile (mmol/L)** |  |  |
| *Total Cholesterol* | 4.8 ± 1.0 | 2.5 – 7.3 |
| *HDL Cholesterol* | 1.6 ± 0.4 | 0.9 – 2.8 |
| *LDL Cholesterol* | 2.7 ± 0.8 | 0 – 4.8 |
| *Triglycerides* | 1.2 ± 0.7 | 0.4 – 5.7 |
| *VLDL Cholesterol* | 0.5 ± 0.3 | 0 – 1.4 |
| **MCV (fL)** | 89.4 ± 4.9 | 70 – 107 |

### Prevalence of Prediabetes and Diabetes

The majority of participants (58%, n = 86) were normoglycemic by all three glycemic measures (HbA_1C_, FPG or OGTT). Considering any positive test as diagnostic, five participants (3%) were diagnosed with DM. All 5 subjects had a HbA_1C_ ≥ 6.5% but only one met the criteria for diabetes by OGTT, and none by FPG (Table 2). Two subjects with DM by HbA_1C_ had completely normal FPG and OGTT. Another 39% (n=58) of subjects did not meet any criteria for diabetes but met at least one criterion for prediabetes (Table 2). Prevalence of prediabetes by HbA_1C_ was 35%, by IGT 8.7% and by IFG 8.1%.

**Table 2:**
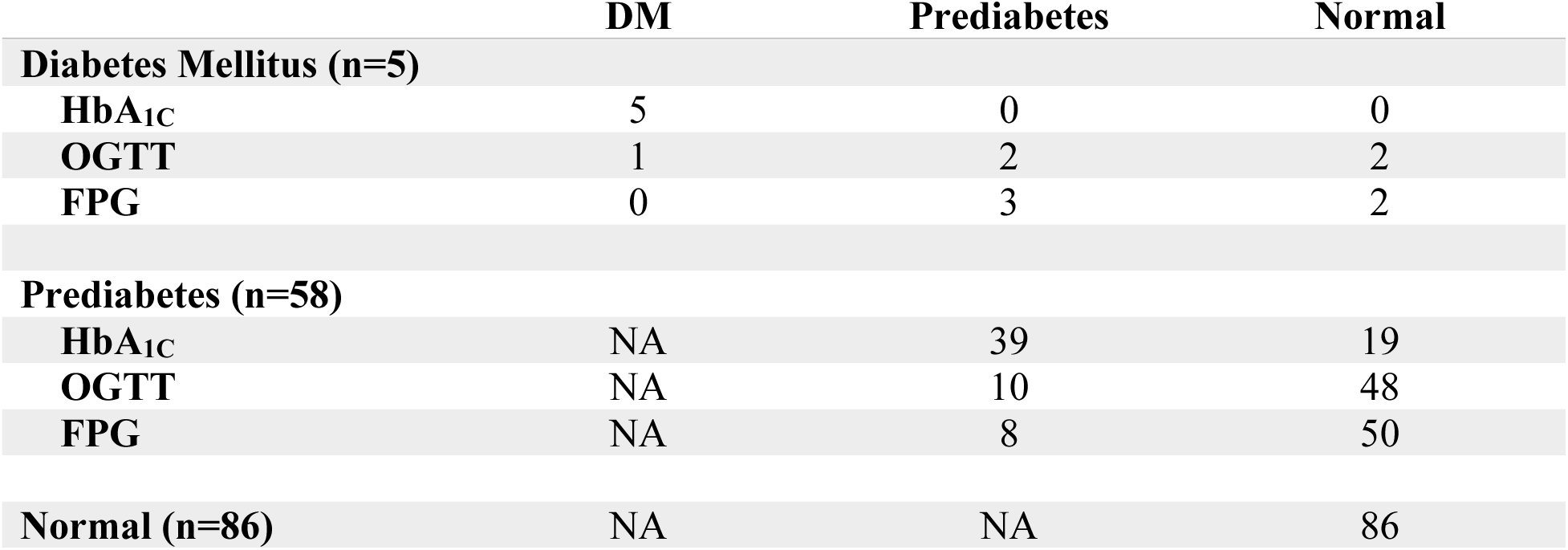
Comparison of glycemic results for Filipino-Americans without diabetes (N = 149). Data are reported as counts.

Focusing on the 58 subjects meeting at least one criterion for prediabetes but no criteria for diabetes, Figure 1 illustrates the overlap between abnormal results from the different tests. Sixty seven percent of the participants with prediabetes met only the HbA_1C_ criteria. Only one subject met prediabetes criteria by all three measures.

**Figure 1:**
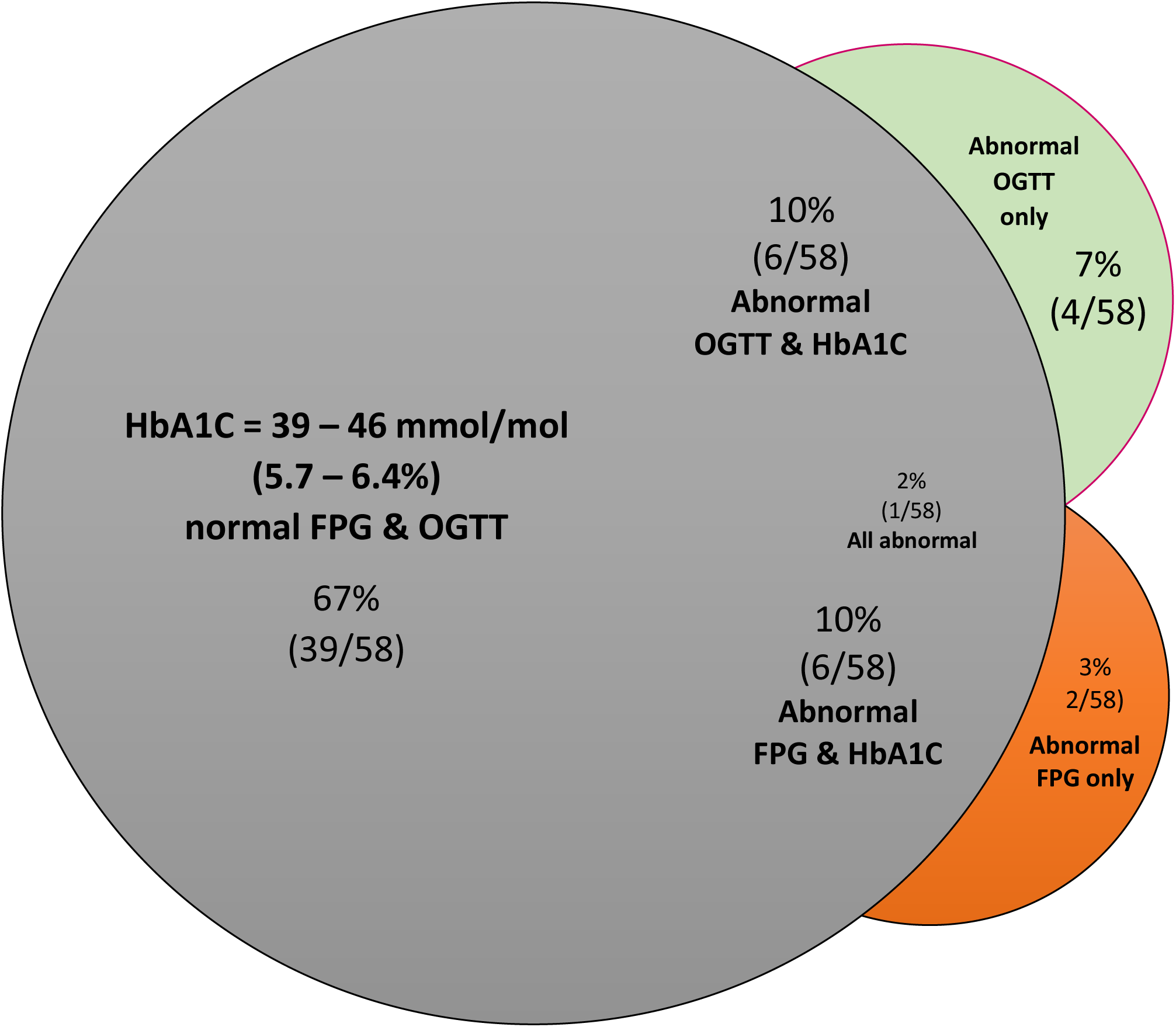
Prediabetes diagnostic concordance using FPG, OGTT and HbA_1C_ in Filipino-American subjects with at least one test meeting the criteria for prediabetes (n = 58). Prediabetes defined as: FPG = 5.6 – 6.9 mmol/L (orange circle), OGTT glucose = 7.8 – 11 mmol/L (green circle), HbA_1C_ = 39 – 46 mmol/mol (5.7 – 6.4%, grey circle).

Given so many subjects met diagnostic criteria for prediabetes or diabetes by HbA_1C_ alone, we wanted to explore whether or not HbA_1C_ was potentially over or under diagnosing diabetes and prediabetes. Therefore, we looked at clinical characteristics in patients in the different A1C diagnostic ranges (Table 3). With increasing HbA_1C_, there was a statistically significant increase in age, BMI, waist-hip ratio, blood pressure (systolic and diastolic), triglycerides, VLDL, FPG and OGTT, insulin and HOMA-IR. The increases in insulin were especially notable. As statin intake may confound our observed difference, we did a subgroup analysis of the non-diabetic individuals not taking statins (n = 128). We found that differences in lipids across HbA_1C_ values were even more significant (Supplementary Table).

**Table 3:** Comparison of characteristics of self-reported Filipino-Americans without diabetes (N = 149) according to glycemic status by HbA_1C_. Data are reported as mean ± standard deviation or n (%), where applicable. BMI - body mass index; FPG – fasting plasma glucose; OGTT – oral glucose tolerance test. *There was only one male with HbA_1C_ ≥ 6.5 and so statistical comparison across the 3 groups was not possible.

|  | HbA<sub>1c</sub> |  |  | p-value |
| --- | --- | --- | --- | --- |
| | < 5.7<br>(n = 92) | 5.7 – 6.4<br>(n = 52) | ≥ 6.5<br>(n = 5) | |
| **Age (years)** | 34 ± 15 | 55 ± 17 | 62 ± 9 | <0.001 |
| **Female** | 55 (60) | 25 (48) | 4 (80) | 0.25 |
| **Positive Family History of DM** | 67 (73) | 30 (58) | 3 (60) | 0.13 |
| **BMI (kg/m<sup>2</sup>)** | 25.0 ± 3.7 | 26.8 ± 3.9 | 27.2 ± 3.1 | 0.02 |
| **Waist-Hip-Ratio** | 0.85 ± 0.07 | 0.90 ± 0.07 | 0.91 ± 0.02 | <0.001 |
| **Waist Circumference (cm)** |  |  |  |  |
| *Female* | 78.5 ± 9.3 | 85.8 ± 12.8 | 90.8 ± 5.2 | <0.01 |
| *Male* | 87.0 ± 10.5 | 93.7 ± 8.3 | 85.0* | NA* |
| **Blood Pressure (mmHg)** |  |  |  |  |
| *Systolic* | 120 ± 17 | 134 ± 18 | 142 ± 16 | <0.001 |
| *Diastolic* | 72 ± 9 | 79 ± 11 | 77 ± 11 | <0.001 |
| **Oral Glucose Tolerance Test (mmol/L)** |  |  |  |  |
| *0 Hour* | 4.6 ± 0.5 | 5.1 ± 0.5 | 5.8 ± 0.7 | <0.001 |
| *2<sup>nd</sup> Hour* | 5.3 ± 1.5 | 5.6 ± 1.8 | 9.2 ± 3.0 | 0.02 |
| **Insulin (pmol/L)** | 59.0 ± 38.2 | 95.2 ± 90.3 | 122.2 ± 50.7 | <0.001 |
| **HOMA-IR** | 1.6 ± 1.2 | 2.9 ± 3.0 | 4.2 ± 2.0 | <0.001 |
| **Lipid Profile (mmol/L)** |  |  |  |  |
| *Total Cholesterol* | 4.7 ± 0.9 | 5.1 ± 1.0 | 4.6 ± 0.6 | 0.05 |
| *HDL Cholesterol* | 1.6 ± 0.4 | 1.5 ± 0.4 | 1.3 ± 0.1 | 0.14 |
| *LDL Cholesterol* | 2.7 ± 0.8 | 2.8 ± 0.9 | 2.6 ± 0.7 | 0.43 |
| *Triglycerides* | 1.0 ± 0.5 | 1.4 ± 0.9 | 1.5 ± 0.7 | <0.001 |
| *VLDL Cholesterol* | 0.5 ± 0.2 | 0.6 ± 0.3 | 0.7 ± 0.3 | <0.01 |

We also compared those with both a normal FPG and OGTT (n=127) to those that had any abnormal FPG or OGTT including the one individual positive for DM by OGTT (n=22) (Table 4), irrespective of HbA_1C_. Individuals with dysglycemia by FPG/OGTT were significantly older and had higher waist-hip ratio (women only), systolic blood pressure, HbA_1C_, insulin and HOMA-IR. There were no differences in lipids.

**Table 4:** Comparison of characteristics of self-reported Filipino-Americans without diabetes (N = 149) according to glycaemic status by normal FPG and OGTT versus dysglycemia (either or both FPG and OGTT abnormal). Data are reported as mean ± standard deviation or n (%), where applicable. BMI - body mass index; FPG – fasting plasma glucose; OGTT – oral glucose tolerance test

|  | **FPG and/or OGTT** |  | **p-value** |
| --- | --- | --- | --- |
|  | **Normal**<br>**(n = 127)** | **Dysglycaemia**<br>**(n = 22)** |  |
| **Age (years)** | 39 ± 17 | 59 ± 19 | <0.001 |
| **Female** | 74 (58) | 10 (45) | 0.35 |
| **Positive Family History of DM** | 87 (69) | 13 (59) | 0.46 |
| **BMI (kg/m<sup>2</sup>)** | 25.5 ± 3.8 | 27.0 ± 3.8 | 0.08 |
| **Waist-Hip-Ratio** | 0.86 ± 0.07 | 0.93 ± 0.07 | <0.001 |
| **Waist Circumference (cm)** |  |  |  |
| *Female* | 79.8 ± 9.5 | 92.1 ± 14.9 | <0.001 |
| *Male* | 89.1 ± 9.7 | 92.8 ± 11.4 | 0.26 |
| **Blood Pressure (mmHg)** |  |  |  |
| *Systolic* | 123 ± 17 | 140 ± 21 | <0.001 |
| *Diastolic* | 74 ± 10 | 79 ± 11 | 0.05 |
| **HbA<sub>1C</sub> (%)** | 5.5 ± 0.4 | 6.0 ± 0.6 | <0.001 |
| *HbA<sub>1C</sub> (mmol/mol)* | 37 ± 4.4 | 42 ± 6.6 | |
| **Insulin (pmol/L)** | 64.6 ± 39.6 | 128.5 ± 127.8 | <0.001 |
| **HOMA-IR** | 1.8 ± 1.2 | 4.2 ± 4.2 | <0.001 |
| **Lipid Profile (mmol/L)** |  |  |  |
| *Total Cholesterol* | 4.8 ± 1.0 | 4.8 ± 1.0 | 0.90 |
| *HDL Cholesterol* | 1.6 ± 0.4 | 1.5 ± 0.4 | 0.46 |
| *LDL Cholesterol* | 2.7 ± 0.9 | 2.7 ± 0.8 | 0.86 |
| *Triglycerides* | 1.17 ± 0.7 | 1.2 ± 0.6 | 0.69 |
| *VLDL Cholesterol* | 0.5 ± 0.3 | 0.6 ± 0.3 | 0.39 |

### Association of HbA_1C_ with glucose tests

The Supplementary Figure shows the relationships of HbA_1C_ with FPG and OGTT glucose. In an unadjusted linear model (Supplementary Figure A) an HbA_1C_ of 39 mmol/mol (5.7%) corresponded to a FPG of 4.9 mmol/L (88 mg/dL). Every 10.9 mmol/mol (1%) increase in HbA_1C_ corresponded to a 0.8 mmol/L (14 mg/dL) (95% CI 0.6 – 0.9 mmol/L, p <0.001) increase in FPG. After adjusting for age, sex and BMI the relationship shifted with a 10.9 mmol/mol (1%) increase in HbA_1C_ corresponding to only a 0.6 mmol/L (10.8 mg/dl) (95% CI 0.4 – 0.8 mmol/L, p < 0.001) increase in FPG.

The unadjusted linear model comparing HbA_1C_ and OGTT glucose (Supplementary Figure B), showed HbA_1C_ of 39 mmol/mol (5.7%) corresponded to an OGTT glucose of 5.4 mmol/L (97.2 mg/dL) and OGTT glucose increased by an average of 0.8 mmol/L (15 mg/dL) (95% CI 0.2 – 1.5 mmol/L, p = 0.02) with every 10.9 mmol/mol (1%) increase in HbA_1C_. After adjusting for BMI, age and gender, every 10.9 mmol/mol (1%) increase in HbA_1C_ corresponded to an average increase of 0.4 mmol/L (8.02 mg/dL) (95% CI -0.4 – 1.3 mmol/L) of OGTT but this did not reach statistical significance (p = 0.28).

## Discussion

In a Filipino-American without previously diagnosed diabetes cohort we found HbA_1C_, FPG and OGTT were highly discordant. Agreement between any combination of the three measures was low. The 35% prevalence of prediabetes by HbA_1C_ was an astounding four-fold greater than that seen with FPG (8.1%) or OGTT (8.7%). Given the concern for overdiagnosis by HbA_1C_, we explored whether metabolic indicators of insulin resistance and the metabolic syndrome were present in this group. Across the HbA_1C_ categories, individuals categorized with prediabetes by HbA_1C_ had statistically significantly higher HOMA-IR, fasting insulin, waist circumference, waist to hip ratio, BMI, blood pressure, triglycerides and VLDL cholesterol compared to their normoglycemic counterparts. This strongly supports the theory that this high prevalence of prediabetes seen with HbA_1C_ alone corresponds to a clinically relevant phenotype – a phenotype suggestive of insulin resistance and on the path to diabetes. Future prospective studies are needed to confirm this hypothesis.

The highly discordant and variable prevalence of prediabetes by HbA_1C_ depending on racial or ethnic group is concerning. Our findings showed a higher prevalence of prediabetes by HbA_1C_ among Filipino-Americans similar to the results among Chinese^13^, high metabolic risk Hispanic^14^, Palestinian Arabs^15^ and South Asians^20^. Filipinos are a complex admixture of Chinese, Native Filipino, Spanish, and Native Central and South American^25^ genetic ancestry with the percent contribution from these groups differing depending on geographic region in the Philippines. Future studies assessing genetic ancestry could reveal whether or not this finding of high prediabetes prevalence by HbA_1C_ is more closely associated with any particular genetic ancestry in Filipinos.

All glycemic measures for prediabetes are surrogate markers for adverse outcomes and not true disease per se. Prospective studies looking at the progression to diabetes in diverse populations (e.g. Japanese^18,19^, Chinese^26^, Finnish^27^, European^28^, multi-racial American^29^) do exist, and they have generally shown that HbA_1C_ in the prediabetes range is comparable to or better than IFG. However, IGT seems to be a superior marker for eventual diabetes^26–28^ in some groups.

It is important to note that the recommendation for detection of prediabetes using HbA_1C_ is more recent compared to FPG and OGTT. When the cut point for IFG was first introduced, a discordance between IFG and IGT led the ADA to adjust criteria for IFG^30^. Perhaps, as data accumulates, consideration should be made for a re-evaluation of the HbA_1C_ diagnostic criteria with a consideration for criteria adjustment depending on the patient’s racial and ethnic groups as appropriate. More prospective studies are needed to guide those potential changes, including in a Filipino population.

Our study has several limitations. First, we relied on cross-sectional data using single measurements of FPG, OGTT and HbA_1C_. This could have resulted in misclassification of diagnosis especially in individuals with values that are near the cut-offs for FPG and OGTT. Second, given that the parent study was not powered for this type of descriptive analysis, our sample size is limiting and may have hampered our ability to find associations. Third, we did not directly assess for hemoglobinopathies. One mechanism proposed by previous studies to explain the discordance in diagnosis by HbA_1C_ is the existence of high rates of hemoglobinopathies^31^ in some populations. It has been reported that in Filipinos, the gene for α-thalassemia has a frequency of 5% while frequency of β-thalassemia has been variously reported from 1 to 9%^32^. We did exclude individuals with MCV < 70 fL from our cohort, however underlying haemoglobinopathies could still exist confounding our results.

In summary, we found a four-fold higher prevalence of prediabetes in a Filipino-American cohort using HbA_1C_ compared to FPG or OGTT. The degree of discordance between measures in this cohort is concerning and has profound implications for prediabetes screening in the Philippines and in expatriate Filipino populations across the world. When faced with similar results, others have recommended against the use of HbA_1C_ for detecting prediabetes^16^. However, as the true gold standard for prediabetes is not known, without prospective studies, that conclusion seems premature and could potentially result in significant underdiagnosis in this population. Our data show individuals diagnosed with prediabetes by HbA_1C_ have metabolic profiles consistent with insulin resistance suggesting those individuals are truly on the path to diabetes. This small study highlights the urgent need for larger studies and outcome studies to determine the true diabetes risk in this and other diverse populations.

## Data Availability

All data produced in the present study are available upon reasonable request to the authors.

## Acknowledgements

We would like to acknowledge San Francisco General Hospital, Philippine General Hospital, and the University of the Philippines Manila – National Institutes of Health for providing the space and facilities to conduct this research study.

This research was supported by a grant from the Philippine-California Advance Research Initiative Program of the Commission on Higher Education (CHED-PCARI) in partnership with the Department of Science and Technology of the Philippines.

Disclosure Summary: Maria Elizabeth Mercado was granted a full scholarship for a Master’s Degree at University of California, San Francisco (UCSF) by CHED-PCARI. Elizabeth J. Murphy was supported by the Deborah Cowan Endowed Professorship. All other authors have no conflict of interest or other source of funding to disclose.

We would also like to acknowledge Dr Neil Risch for his insights and contributions to the ideas that formed our understanding of the data and Dr Peter Bacchetti for his guidance in the statistical analysis. Lastly, we would like to acknowledge Dr. Margarette Mariano, Dr. Alvin Lirio and the recruitment staff who oversaw the collection and day to day operations of the Filipino Health Study.

---

**Supplementary Table:** Comparison of lipid profiles of Filipino-Americans without diabetes mellitus not using any statin drug classified as normal and with prediabetes based on OGTT HbA1C. Of note all patients with diabetes were on a statin and therefore are not included. Data are reported as mean ± standard deviation

|  | HbA1C |  | p-value |
| --- | --- | --- | --- |
|  | < 5.7<br>(n = 90) | 5.7 – 6.4<br>(n = 38) |  |
| Lipid Profile (mmol/L) |  |  |  |
| *Total Cholesterol* | 4.7 ± 1.0 | 5.3 ± 0.8 | < 0.001 |
| *HDL Cholesterol* | 1.6 ± 0.4 | 1.6 ± 0.4 | 0.79 |
| *LDL Cholesterol* | 2.7 ± 0.8 | 3.1 ± 0.7 | < 0.01 |
| *Triglycerides* | 1.0 ± 0.5 | 1.4 ± 0.7 | <0.001 |
| *VLDL Cholesterol* | 0.5 ± 0.2 | 0.6 ± 0.3 | <0.001 |

**Supplementary Figure:**
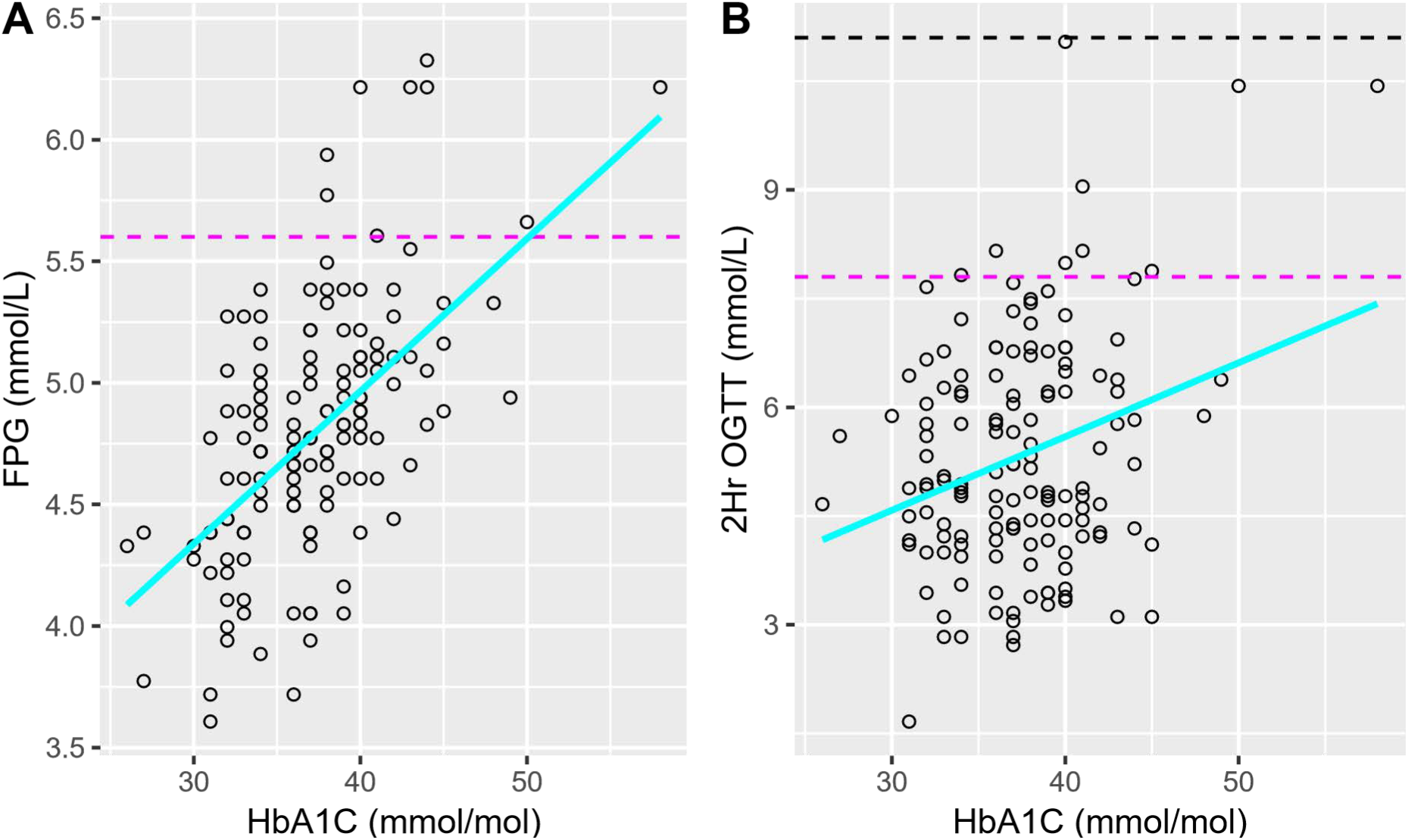
Scatter plot of the relationship between (A) HbA_1C_ and FPG (n = 149) and (B) HbA_1C_ and OGTT (n = 143). Linear estimate (cyan) with 95% confidence intervals (shaded area); --- Cut-off for prediabetes diagnosis based on corresponding glucose test; --- Cut-off for diabetes mellitus diagnosis based on corresponding glucose test.

## References

1. American Diabetes Association. Standards of Medical Care in Diabetes—2010. Diabetes Care. 2010;33(Supplement 1):S11–S61. doi:10.2337/dc10-S011

2. Dagogo-Jack S. Pitfalls in the use of HbA1c as a diagnostic test: the ethnic conundrum. Nat Rev Endocrinol. 2010;6(10):589–593. doi:10.1038/nrendo.2010.126

3. Radin MS. Pitfalls in Hemoglobin A1c Measurement: When Results may be Misleading. J Gen Intern Med. 2014;29(2):388–394. doi:10.1007/s11606-013-2595-x

4. Christensen DL, Witte DR, Kaduka L, Jørgensen ME, Borch-Johnsen K, Mohan V, Shaw JE, Tabák AG, Vistisen D. Moving to an A1C-Based Diagnosis of Diabetes Has a Different Impact on Prevalence in Different Ethnic Groups. Diabetes Care. 2010;33(3):580–582. doi:10.2337/dc09-1843

5. Yip WCY, Sequeira IR, Plank LD, Poppitt SD. Prevalence of Pre-Diabetes across Ethnicities: A Review of Impaired Fasting Glucose (IFG) and Impaired Glucose Tolerance (IGT) for Classification of Dysglycaemia. Nutrients. 2017;9(11):1273. doi:10.3390/nu9111273

6. Sequeira IR, Poppitt SD. HbA1c as a marker of prediabetes: A reliable screening tool or not? Insights Nutr Metab. 2017;1(1):21–29.

7. Guo F, Moellering DR, Garvey WT. Use of HbA1c for Diagnoses of Diabetes and Prediabetes: Comparison with Diagnoses Based on Fasting and 2-Hr Glucose Values and Effects of Gender, Race, and Age. Metab Syndr Relat Disord. 2014;12(5):258–268. doi:10.1089/met.2013.0128

8. Ziemer DC. Glucose-Independent, Black–White Differences in Hemoglobin A _1c_ Levels: A Cross-sectional Analysis of 2 Studies. Ann Intern Med. 2010;152(12):770. doi:10.7326/0003-4819-152-12-201006150-00004

9. Herman WH, Dungan KM, Wolffenbuttel BHR, Buse JB, Fahrbach JL, Jiang H, Martin S. Racial and Ethnic Differences in Mean Plasma Glucose, Hemoglobin A1c, and 1,5-Anhydroglucitol in Over 2000 Patients with Type 2 Diabetes. J Clin Endocrinol Metab. 2009;94(5):1689-1694. doi:10.1210/jc.2008-1940

10. Herman WH, Ma Y, Uwaifo G, Haffner S, Kahn SE, Horton ES, Lachin JM, Montez MG, Brenneman T, Barrett-Connor E. Differences in A1C by Race and Ethnicity Among Patients With Impaired Glucose Tolerance in the Diabetes Prevention Program. Diabetes Care. 2007;30(10):2453–2457. doi:10.2337/dc06-2003

11. Use of Glycated Haemoglobin (HbA1c) in the Diagnosis of Diabetes Mellitus: Abbreviated Report of a WHO Consultation. World Health Organization; 2011:1-25. Accessed June 12, 2018. http://linkinghub.elsevier.com/retrieve/pii/S0168822711001318

12. American Diabetes Association. 6. Glycemic Targets: Standards of Medical Care in Diabetes—2018. Diabetes Care. 2018;41(Supplement 1):S55-S64. doi:10.2337/dc18-S006

13. Li J, Ma H, Na L, Jiang S, Lv L, Li G, Zhang W, Na G, Li Y, Sun C. Increased Hemoglobin A1c Threshold for Prediabetes Remarkably Improving the Agreement Between A1c and Oral Glucose Tolerance Test Criteria in Obese Population. J Clin Endocrinol Metab. 2015;100(5):1997–2005. doi:10.1210/jc.2014-4139

14. Vega-Vázquez MA, Ramírez-Vick M, Muñoz-Torres FJ, González-Rodríguez LA, Joshipura K. Comparing Glucose and Hemoglobin A1C Diagnostic Tests Among A High Metabolic Risk Hispanic Population. Diabetes Metab Res Rev. 2017;33(4). doi:10.1002/dmrr.2874

15. Kharroubi AT, Darwish HM, Abu Al-Halaweh AI, Khammash UM. Evaluation of Glycated Hemoglobin (HbA1c) for Diagnosing Type 2 Diabetes and Prediabetes among Palestinian Arab Population. PLoS ONE. 2014;9(2). doi:10.1371/journal.pone.0088123

16. Shimodaira M, Okaniwa S, Hanyu N, Nakayama T. Optimal Hemoglobin A1c Levels for Screening of Diabetes and Prediabetes in the Japanese Population. J Diabetes Res. Published online 2015. doi:10.1155/2015/932057

17. Booth RA, Jiang Y, Morrison H, Orpana H, Rogers Van Katwyk S, Lemieux C. Ethnic dependent differences in diagnostic accuracy of glycated hemoglobin (HbA1c) in Canadian adults. Diabetes Res Clin Pract. 2018;136:143–149. doi:10.1016/j.diabres.2017.11.035

18. Heianza Y, Hara S, Arase Y, Saito K, Fujiwara K, Tsuji H, Kodama S, Hsieh SD, Mori Y, Shimano H, Yamada N, Kosaka K, Sone H. HbA1c 5·7–6·4% and impaired fasting plasma glucose for diagnosis of prediabetes and risk of progression to diabetes in Japan (TOPICS 3): a longitudinal cohort study. The Lancet. 2011;378(9786):147–155. doi:10.1016/S0140-6736(11)60472-8

19. Heianza Y, Arase Y, Fujihara K, Tsuji H, Saito K, Hsieh SD, Kodama S, Shimano H, Yamada N, Hara S, Sone H. Screening for pre-diabetes to predict future diabetes using various cut-off points for HbA1c and impaired fasting glucose: the Toranomon Hospital Health Management Center Study 4 (TOPICS 4). Diabet Med. 2012;29(9):e279–e285. doi:10.1111/j.1464-5491.2012.03686.x

20. Bhowmik B, Diep LM, Munir SB, Rahman M, Wright E, Mahmood S, Afsana F, Ahmed T, Khan AKA, Hussain A. HbA1c as a diagnostic tool for diabetes and pre-diabetes: the Bangladesh experience. Diabet Med. 2013;30(3):e70–e77. doi:10.1111/dme.12088

21. Kramer CK, Araneta MRG, Barrett-Connor E. A1C and Diabetes Diagnosis: The Rancho Bernardo Study. Diabetes Care. 2010;33(1):101–103. doi:10.2337/dc09-1366

22. Araneta MRG, Grandinetti A, Chang HK. A1C and Diabetes Diagnosis Among Filipino Americans, Japanese Americans, and Native Hawaiians. Diabetes Care. 2010;33(12):2626–2628. doi:10.2337/dc10-0958

23. World Health Organization. Waist Circumference and Waist-Hip Ratio: Report of a WHO Expert Consultation, Geneva, 8-11 December 2008. World Health Organization; 2011.

24. Matthews DR, Hosker JP, Rudenski AS, Naylor BA, Treacher DF, Turner RC. Homeostasis model assessment: insulin resistance and β-cell function from fasting plasma glucose and insulin concentrations in man. Diabetologia. 1985;28(7):412–419. doi:10.1007/BF00280883

25. Banda Y, Kvale MN, Hoffmann TJ, Hesselson SE, Ranatunga D, Tang H, Sabatti C, Croen LA, Dispensa BP, Henderson M, Iribarren C, Jorgenson E, Kushi LH, Ludwig D, Olberg D, Quesenberry CP, Rowell S, Sadler M, Sakoda LC, Sciortino S, Shen L, Smethurst D, Somkin CP, Eeden SKVD, Walter L, Whitmer RA, Kwok PY, Schaefer C, Risch N. Characterizing Race/Ethnicity and Genetic Ancestry for 100,000 Subjects in the Genetic Epidemiology Research on Adult Health and Aging (GERA) Cohort. Genetics. 2015;200(4):1285–1295. doi:10.1534/genetics.115.178616

26. Lu J, He J, Li M, Tang X, Hu R, Shi L, Su Q, Peng K, Xu M, Xu Y, Chen Y, Yu X, Yan L, Wang T, Zhao Z, Qin G, Wan Q, Chen G, Dai M, Zhang D, Gao Z, Wang G, Shen F, Luo Z, Qin Y, Chen L, Huo Y, Li Q, Ye Z, Zhang Y, Du R, Cheng D, Liu C, Wang Y, Wu S, Yang T, Deng H, Li D, Lai S, Bloomgarden ZT, Chen L, Zhao J, Mu Y, Ning G, Wang W, Bi Y, for the 4C Study Group. Predictive Values of Fasting Glucose, Postload Glucose, and Hemoglobin A1c on Risk of Diabetes and Complications in Chinese Adults. Diabetes Care. 2019;42:1539-1548. doi:10.2337/dc18-1390

27. Cederberg H, Saukkonen T, Laakso M, Jokelainen J, Harkonen P, Timonen M, Keinanen-Kiukaanniemi S, Rajala U. Postchallenge Glucose, A1C, and Fasting Glucose as Predictors of Type 2 Diabetes and Cardiovascular Disease: A 10-year prospective cohort study. Diabetes Care. 2010;33(9):2077–2083. doi:10.2337/dc10-0262

28. Shahim B, De Bacquer D, De Backer G, Gyberg V, Kotseva K, Mellbin L, Schnell O, Tuomilehto J, Wood D, Rydén L. The Prognostic Value of Fasting Plasma Glucose, Two-Hour Postload Glucose, and HbA _1c_ in Patients With Coronary Artery Disease: A Report From EUROASPIRE IV: A Survey From the European Society of Cardiology. Diabetes Care. 2017;40(9):1233-1240. doi:10.2337/dc17-0245

29. Selvin E, Matsushita K, Pankow J, Brancati FL. Glycated Hemoglobin, Diabetes, and Cardiovascular Risk in Nondiabetic Adults. N Engl J Med. Published online 2010:12.

30. The Expert Committee on the Diagnosis and Classification of Diabetes. Follow-up Report on the Diagnosis of Diabetes Mellitus. Diabetes Care. 2003;26(11):3160-3167. doi:10.2337/diacare.26.11.3160

31. Sacks DB. A1C Versus Glucose Testing: A Comparison. Diabetes Care. 2011;34(2):518–523. doi:10.2337/dc10-1546

32. Fucharoen S, Winichagoon P. Haemoglobinopathies in Southeast Asia. Indian J Med Res. 2011;134(4):498–506.

